# How do Structural Barriers to Green and Blue Spaces Influence the Prescription of Nature-Based Interventions: A Scoping Review Protocol

**DOI:** 10.1101/2020.07.03.20145946

**Authors:** Rachel M. Nejade, Daniel M. Grace, Leigh R. Bowman

## Abstract

**Introduction:** Emerging evidence has demonstrated that nature-based interventions (NBIs) can improve mental and physical health. Considering that the global burden of poor mental health continues to rise, such interventions could be a cost-effective means to improve mental health, as well as reconnect individuals with the natural world, and thus aid conservation efforts. However, the effectiveness of NBIs as a prescriptive intervention is, in part, a function of access to blue and green spaces. Accordingly, this scoping review will explore how structural inequalities influence the effectiveness of nature-based interventions as treatment options for mental and physical ill health.

**Methods and Analysis:** A scoping review will be conducted to identify the barriers and facilitators associated with the utilisation of green and blue spaces. The review will follow the PRISMA-ScR guidelines, in addition to the associated Cochrane guidelines for scoping reviews. A literature search will be performed across five databases, and articles will be selected based on key inclusion/ exclusion criteria. All data will be extracted to a pre-defined charting table. The primary and secondary outcomes will be mental and physical health respectively.

**Ethics and Dissemination:** All data rely on secondary, publicly available data sources; therefore no ethical clearance is required.

Upon completion, the results of this study will be disseminated via the Imperial College London Community and published in an open access, peer-reviewed journal.

**Article Summary:** *Strengths and Limitations of this Study:*

- This scoping review protocol is the first to focus on the accessibility to green and blue spaces in the context of mental and physical health.
- This protocol and subsequent review benefit from increased transparency, a systematised strategy (PRISMA-ScR), and a reduction in the risk of bias, through publication in an open access journal.
- This review will also capture grey literature - studies published outside peer-reviewed journals.
- Due to the broad nature of the review, the research may unearth more questions than solutions.

*Registration Number:* Open Science Framework: 10.17605/OSF.IO/8J5Q3

## Introduction

In 2017, 792 million people worldwide lived with a mental health disorder (1), defined as “syndromes characterized by clinically significant disturbance in an individual’s cognition, emotional regulation, or behaviour that reflects a dysfunction in the psychological, biological, or developmental processes that underlie mental and behavioural functioning” (ICD-11, 2020) (2). This growing prevalence of poor mental health has both direct and indirect costs, and in financial terms, is estimated at between £70-100 billion annually for the United Kingdom (UK) alone (3).

A growing body of evidence suggests that nature-based interventions (NBIs) could be a cost-effective solution to improve mental health outcomes, while also addressing the increasing demand from patients for less intrusive treatment options (4). Indeed, contact with nature has been shown to reduce stress and improve self-reported emotions, as well as increase attention capacity, happiness, and vitality (5, 6, 7). It has also been shown to improve physiological outcomes such as longevity and self-reported health (8).

Over time, NBIs have been referred to as ‘green care’, ‘ecotherapy’, ‘social prescribing’ or ‘green prescribing’, demonstrating the lack of consensus on terminology in this field (6). ‘Blue prescribing’ is also increasingly considered a part of NBIs, since water environments (e.g. river, coastlines) are as much a part of nature as green spaces (e.g. parks, public open spaces) (9).

A recent technical report by Robinson (2018) defines green prescribing as a “non-medical intervention designed to improve physical and mental health through exposure to and interaction with natural environments”, by prescribing various monitorable activities such as nature walks, physical exercise, care farming or horticulture (10). Bragg and Atkins (2016) have offered definitions for a range of NBI activities in relation to green spaces (Supplementary File: Annex A_Types of NBIs) (6). Although not necessarily included within Robinson’s (2018) definition, blue prescribing remains very similar in theory, where the activities prescribed refer instead to physical activities such as swimming (11).

### Rationale

Given the positive if limited evidence base, NBIs are now prescribed to patients in New Zealand and Scotland, as a means to improve mental health. While this is a positive step, NBIs nonetheless rely on patients being able to access green and blue spaces (12). Yet, environmental (i.e. quality, land management) and socio-structural barriers (i.e. socio-economic status, public transport infrastructure etc.) to green and blue spaces limit the accessibility of NBIs in both rural and urban areas (12). Consequently, a scoping review will be conducted to assess how the barriers to the utilisation of green and blue spaces influence the effectiveness of nature-based interventions.

#### Objectives

This review will be based around the following three objectives:

1. To locate and review the evidence base for green and blue interventions for mental and physical health outcomes in the existing literature.
2. To identify the enablers and barriers to green and blue spaces in relation to nature-based interventions for mental and physical health.
3. To understand how these enablers and barriers impact the effectiveness of green and blue spaces on mental and physical health outcomes.

## Methods and Analysis

### Study Design

A scoping review is the most appropriate methodology for this research, due to the capacity to answer broad questions and summarise findings from a heterogeneous body of knowledge. This scoping review will draw on existing literature to describe the evidence base for nature-based interventions in mental and physical health, and highlight potential barriers and facilitators associated with the utilisation of green and blue spaces, which may translate into NBI effectiveness.

This review will be conducted in accordance with the Preferred Reporting Items for Systematic Reviews and Meta-Analyses Extension for Scoping Reviews (PRISMA – ScR) guidelines, with consideration given to the associated Cochrane guidelines for scoping reviews.

### Search Strategy

The systematic search strategy was developed by three researchers and will cover terms related to nature-based interventions including ‘green’ and ‘blue’ interventions, mental and physical health outcomes, determinants of health, and of green and blue spaces utilisation. The primary outcome will be mental health (determined using a range of quantitative indicators); the secondary outcome will focus on physical health using a range of parameters. The full list of all the key words used for the literature search can be found in Figure 1. The terminology in the literature search reflects the varied positions held by health professionals on the definition of nature-based interventions and the lack of consensus surrounding their application. This systematic search strategy will include free text and subject heading terms, adapted to each of the databases.

**Figure 1:**
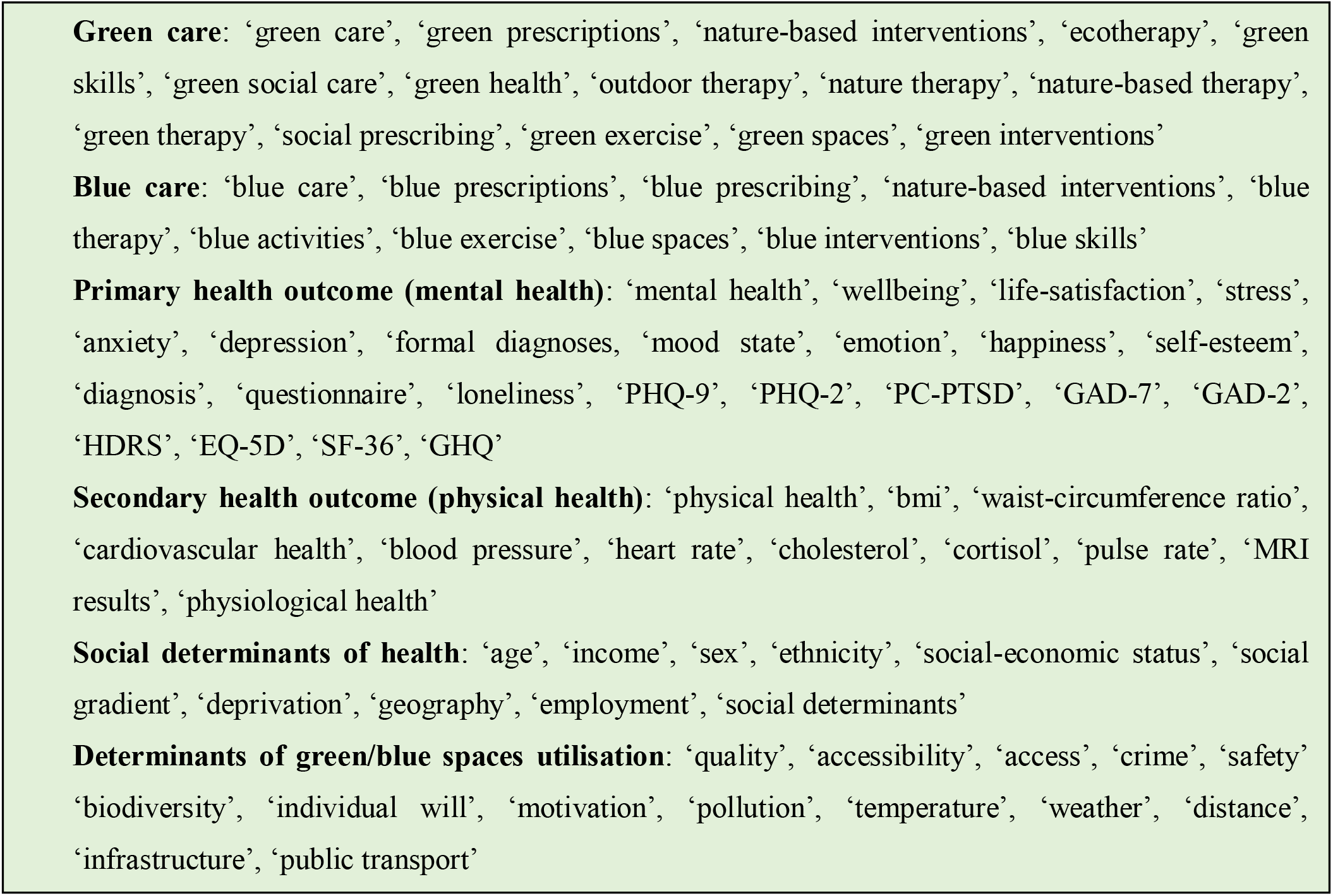
Key terms for the literature search.

Studies will be reviewed across the following five databases: PubMed, The Cochrane Library, OVID (including: Embase, PsycINFO, Global Health, MEDLINE, Health Management Information Consortium (HMIC), Transport Database), Web of Science and Scopus. A search of reference lists from included articles will be performed. The search will also be extended to grey literature using Google Scholar, governmental and institutional websites (i.e. Public Health England (PHE), National Health Service (NHS)). Mendeley and Covidence software will be used to store, organise, and manage all references. An example search strategy, based on the database PubMed, is available in Supplementary file: Annex B_Full Search Strategy, to aid transparency and reproducibility of the results. This search strategies will be adapted for other databases.

### Study Selection Criteria

RN will run all searches, with the support of DG and LB. First stage screening will comprise a review of titles and abstracts in accordance with the inclusion and exclusion criteria. Selected studies will undergo a second stage of screening against the same inclusion/ exclusion criteria by reviewing the full text. Consensus among the three researchers will be reached where a study is deemed to sit on the border between inclusion and exclusion. Duplicates will be removed from the search at both screening stages.

The inclusion criteria for this scoping review are intentionally broad since this is an emerging field, and the authors anticipate few studies, both narrow and broad in scope. To be included, studies must can be either human-centric or review articles with reference to green or blue spaces and a health perspective. Considering the contemporary topic of this scoping review, only studies from 1980 onwards will be included. Studies must also be published in either French of English due to the absence of additional translational capacity.

Excluded studies will be those that do not fit the inclusion criteria and only report on either NBI exposure or health outcomes alone. Studies will also be excluded if they were conducted before 1980. This should ensure that all relevant studies to the research question are included and analysed accordingly, as well as a number of literature types described in Table 1.

**Table 1:** Inclusion and exclusion criteria for the scoping review.

| Inclusion Criteria | Exclusion Criteria |
| --- | --- |
| ▪ All studies and reviews that report both any NBI and physical or mental health outcome of interest, including:<br>· Peer-reviewed articles (i.e. original research and review articles)<br>· Conference proceedings, dissertations/theses and abstracts published in peer-reviewed journals<br>· Grey literature: Google Scholar and governmental and institutional websites (i.e. NHS, PHE), local health authority publications<br>▪ Published from year 1980 onwards<br>▪ Any study designs<br>▪ Language: English and French | ▪ All studies that report only NBI exposure without physical or mental health outcomes.<br>▪ Literature and study designs including<br>· Books and book chapters<br>· Book reviews<br>· Websites and newspaper articles<br>· Social Media content<br>· Commentaries and opinion papers<br>· Data not in public domain<br>· Unpublished data<br>· Protocols<br>▪ Published before 1980<br>▪ Published in any other language than English or French. |

### Data extraction and analysis

Data extraction (or charting) is a common method used in scoping reviews to extract relevant information from the included sources of evidence. It will be conducted by one researcher using a standardised data extraction form, adapted for this scoping review. A preliminary charting template used to address the research questions and objectives is presented in Table 2. Data extraction will be reviewed by the research group to ensure the cohesion and acceptability of the charting process.

**Table 2:**
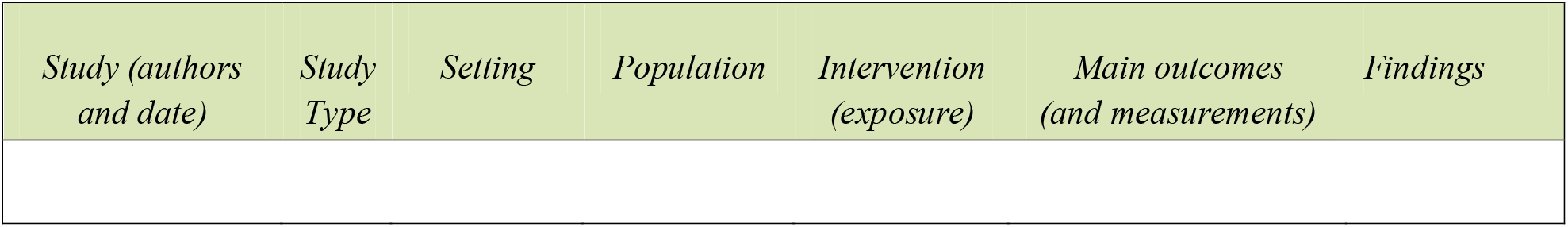
data extraction form (for original studies)

Thematic content analysis will be performed to group findings in categories sharing similarities. This will be performed by one researcher and reviewed by the research group. Any amendments to this scoping review protocol will be documented with reference to saved searches and analysis.

### Patient and Public Involvement

Patients, nor the public, will be involved at any stage during this research.

## Discussion

This review will provide the necessary evidence to inform public health policy, urban planning and primary care on the structural barriers to accessing green and blue spaces. In so doing, key stakeholders will be better informed to develop new policy to mitigate any barriers highlighted by this forthcoming review.

## Data Availability

This protocol has been registered at the following: Open Science Framework: 10.17605/OSF.IO/8J5Q3. All data are available within the manuscript.

## Acknowledgements

The authors would like to thank the School of Public Health at ICL for their continued support.

## Declarations

### Consent for publication

Not Applicable.

### Author contributions

LB conceived this scoping review; LB, DG and RN collaborated in developing the aims and objectives, search strategy, and in selecting which methods to use to conduct this review.

